# Saliva and Blood Cell-Free mtDNA Reactivity to Acute Psychosocial Stress

**DOI:** 10.1101/2025.04.09.25325473

**Authors:** Caroline Trumpff, David Shire, Jeremy Michelson, Natalia Bobba-Alves, Temmie Yu, Richard P. Sloan, Robert-Paul Juster, Michio Hirano, Martin Picard

## Abstract

Human blood contains cell-free mitochondrial DNA (cf-mtDNA) that dynamically increases in concentration in response to acute mental stress. Like other neuroendocrine stress markers, we previously found that cf-mtDNA is also detectable in saliva, calling for studies examining saliva cf-mtDNA reactivity to mental stress. In healthy women and men from the MiSBIE (Mitochondrial Stress, Brain Imaging, and Epigenetics) study (n=68, 66% women), a brief socio-evaluative stressor induced a striking 280% or 2.8-fold increase in saliva cf-mtDNA concentration within 10 minutes (g=0.55, p<0.0001). In blood drawn concurrently with saliva sampling, stress increased cf-mtDNA by an average 32% at 60 min in serum (g=0.20), but not in anticoagulated plasma where cf-mtDNA decreased by 19% at 60 min (g=0.25). Examining the influence of mitochondrial health on cf-mtDNA reactivity in participants with rare mitochondrial diseases (MitoD), we report that a subset of MitoD participants exhibit markedly blunted saliva cf-mtDNA stress reactivity, suggesting that bioenergetic defects within mitochondria may influence the magnitude of saliva, and possibly blood cf-mtDNA responses. Our results document robust saliva cf-mtDNA stress reactivity and provide a methodology to examine the psychobiological regulation of cell-free mitochondria in future studies.

## 1. Introduction

Psychosocial stressors trigger rapid neuroendocrine, metabolic, and immune responses which evolved to mobilize energetic resources required for adaptation, therefore promoting survival ^1^. Over time, chronic stress leads to increased disease risk ^2^ but the physiological pathways for the stress-disease cascade remain unclear ^3^. A recently discovered stress marker is cell-free mitochondrial DNA (cf-mtDNA): fragments or entire mitochondrial genomes that exist in “free” form in the circulation, outside of cells but packaged in different membranous containers ^4–9^. Two studies previously found that plasma and serum cf-mtDNA increase 5-30 minutes after a socio-evaluative challenge ^10,11^. This response pattern is reminiscent of the rapid inducibility of canonical stress hormones, such as cortisol and other acute stress mediators ^12^.

Several stress mediators are detectable not only in blood, but also in saliva ^13^. This led to the discovery that cf-mtDNA exists at high abundance in human saliva ^14^. As it is the case for cortisol, we showed that saliva cf-mtDNA exhibits a robust awakening response, increasing by 2-3-fold within 30-45 minutes after waking up ^14^. Both acute mental stress and waking up in the morning engage energetically costly processes that result in a rise in whole-body energy demand ^15–17^. Mitochondria play an important role in the production of the cellular energetic currency (Adenosine Triphosphate, ATP) and signals required for adaptation ^18^, and it has been suggested that circulating *whole* mitochondria may contribute to processes of resilience and healing ^8,19^, which may support allostasis and adaptive stress responses – all of which consume energy ^20^.

Thus, the stress reactivity of *blood* cf-mtDNA ^10,11,21^ and the existence of cf-mtDNA ^14,22^ in saliva naturally motivated the hypothesis that cell-free mitochondria or naked cf-mtDNA in various biofluids could be dynamically induced by acute psychobiological demands. A recent study investigated this question and found that exposure to a modified Trier social stress test (TSST) caused a 50% increase (trend, not statistically significant) in saliva cf-mtDNA ^21^. Here we examine this question with an 8-timepoint saliva + blood sampling protocol following a brief 5-minute socio-evaluative stress test, using a validated biofluid processing and cf-mtDNA quantification protocol. We demonstrate in healthy individuals and individuals with mitochondrial diseases (MitoD) that saliva cf-mtDNA exhibits rapid and robust reactivity to socio-evaluative stress, well above blood changes. These findings suggest that saliva cf-mtDNA could serve as a bona fide marker of acute mental stress.

## 2. Results

### 2.1. Cf-mtDNA across biofluids

To examine cf-mtDNA dynamics in human blood and saliva in parallel, 68 healthy MiSBIE participants (Mitochondrial Stress, Brain Imaging, and Epigenetics study, 66% women, mean age 37 years) performed a standardized 5-minute socio-evaluative challenge adapted from TSST ^3^. For each participant, we simultaneously collected blood (plasma and serum, intravenous catheter) and saliva (salivette) 5 minutes before and 5, 10, 20, 30, 60, 90, and 120 minutes after the onset of the stressor, as described in ^3^. Biofluids were immediately processed to isolate the cell-free fraction of each sample, and quantify cf-mtDNA using a real-time qPCR-based method (Mitochondrial DNA Quantification by Lysis; MitoQuicLy) ^22^. Previous work showed that plasma and serum contain distinct cf-mtDNA levels ^23,24^ and subtypes of cf-mtDNA particles ^22^, calling for separate analysis of blood-derived plasma (EDTA) and serum (red top), in parallel with our primary biofluid of interest, saliva.

We first compared the amount of cf-mtDNA detected at baseline across biofluids in healthy controls (Figure 1A, B). The average baseline saliva cf-mtDNA concentrations was 1,208 copies per µl. This value was in in the same order of magnitude as blood. On average, saliva levels were 71.4% lower than plasma (p<0.0001, 4,223 copies per µl) and 57.3% lower than serum (p<0.01, 2,831 copies per µl; Figure 1B). Baseline serum and plasma cf-mtDNA levels were not significantly different from one another (p=0.21, Figure 1B).

**Figure 1.**
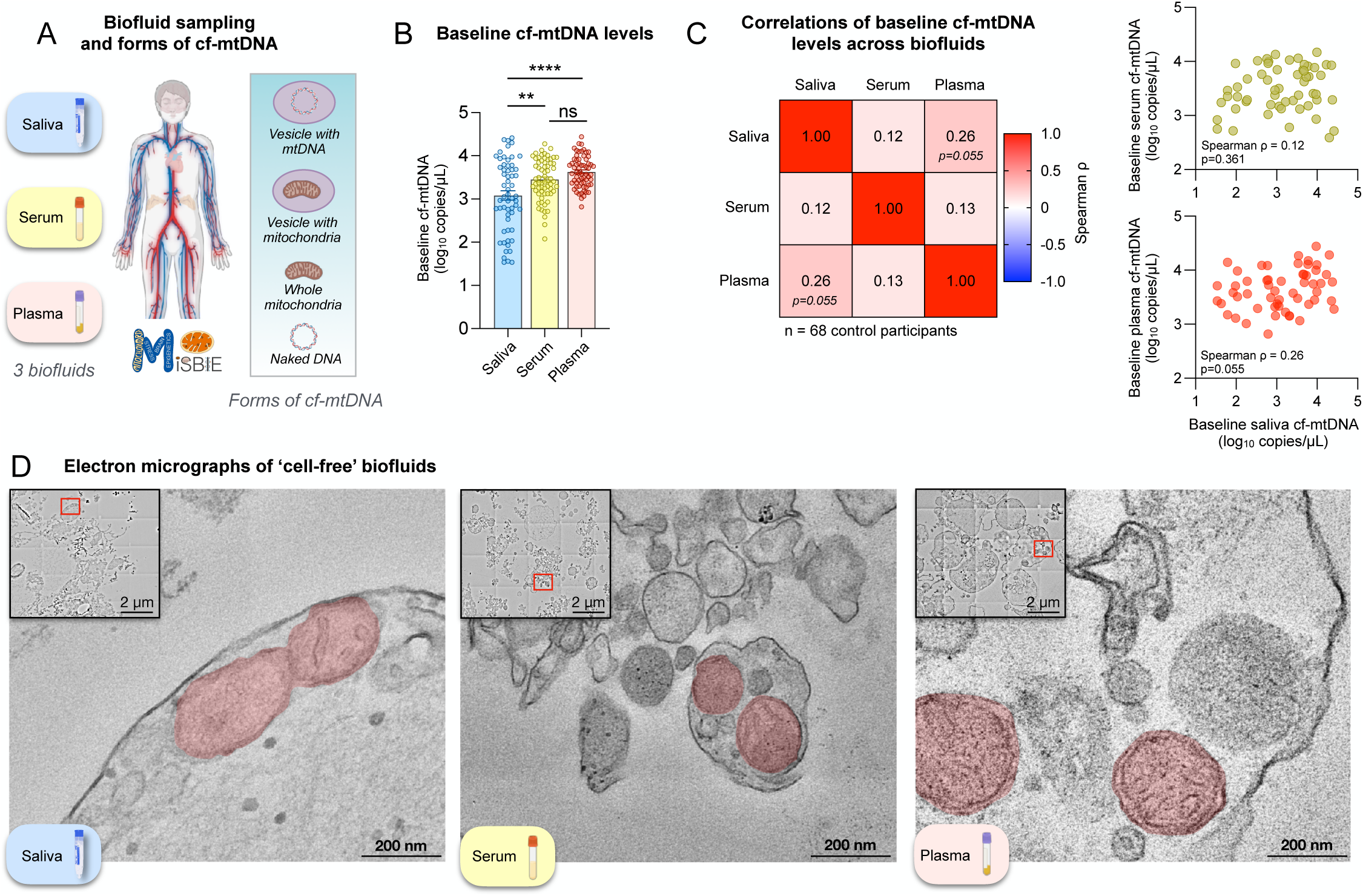
Cell-free mitochondrial DNA in different biofluids. **(A)** Cell-free mitochondrial DNA (cf-mtDNA) is detectable in different biofluids including saliva and blood (serum and plasma) and can be naked or contained within different circulating bodies. **(B)** Comparison of baseline cf-mtDNA levels in saliva, serum, and plasma in control participants (n=59-65). **(C)** Associations between baseline cf-mtDNA levels in saliva (n=59), serum (n=65), and plasma (n=64) shown in heatmap and scatter plots. **(D)** Scanning electron micrographs of membrane-encapsulated structures in saliva, serum, and plasma. Samples used for EM were pre-processed to be rendered ‘cell-free’ following the same protocol used in preparing samples for cf-mtDNA quantification. Red rectangles in insets indicate areas shown in higher magnification images. Red-shading highlights apparent double-membrane structures. Effect sizes and p-values from **(B)** one-way ANOVA with Tukey’s multiple comparisons test and **(C)** Spearman rank correlation. *p<0.05, **p<0.01, ***p<0.001, ****p<0.0001.

Saliva baseline cf-mtDNA levels showed a weak and almost-significant positive correlation with plasma (r=0.260, p=0.055, Figure 1C) but not with serum (r=0.124, p=0.361) cf-mtDNA levels. Consistent with this differential association, plasma and serum cf-mtDNA levels did not show a correlation between each other (r=0.125, p=0.325), illustrating their independent regulation.

cf-mtDNA can be released in the circulation via both active release mechanisms (e.g. vesicles) and possibly cell death. To evaluate the potential role of cell death in the release of cf-mtDNA, we measured another type of cell-free DNA that is known to be released in this process, cell-free nuclear DNA (cf-nDNA) ^9,25–28^. In contrast to cf-mtDNA, baseline saliva cf-nDNA levels were significantly higher than both serum (+204%, p<0.0001) and plasma levels (undetectable in ∼99% of samples) (Figure S1A). cf-mtDNA and cf-nDNA measurements were positively correlated in saliva (r=0.84, p<0.0001; Figure S1C), and to a lesser degree in serum (r=0.40, p<0.0001; Figure S1D). Overall, these findings support the notion that both genomes may be co-released, possibly driven by cell death or other unknown factors, and may therefore contribute to a fraction of cf-mtDNA measured in serum and, to a greater extent, in saliva.

### 2.2. cf-mtDNA biology in plasma, serum and saliva

To examine the biological nature of cf-mtDNA – whether it exists as naked free-floating mtDNA fragments or as membrane bound particles – we performed differential centrifugation experiments where membrane-bound particles are pulled down. In these experiments, free DNA pieces remain in suspension to be detected by qPCR, while membranous vesicular structures (mitochondria, microvesicles) fall to the bottom. These experiments confirmed that the majority (>90%) of cf-mtDNA in human biofluids precipitates when centrifuged at moderate speeds, as we had previously reported ^22^. This means that rather than only naked DNA fragments, the majority of mtDNA detected likely represents sedimentable membranous bodies, such as microvesicles, whole mitochondria, or fragments of mitochondria ^19^.

To confirm this technical point and gain initial insight into the above-described results, we performed electron microscopy imaging studies of mtDNA-enriched fractions from human saliva, serum and plasma from 10 MiSBIE participants. As expected from previous findings ^4,22^, each biofluid contained morphologically distinct biological entities ranging from small vesicles of varying electron density (i.e., containing distinct molecular cargos) to large, heterogenous multivesicular bodies, including several double-membrane structures consistent with swollen or distended mitochondria (Figure 1D). Thus, together with other work ^4,19,22^, these ultrastructural observations from mtDNA-rich saliva, serum, and plasma fractions demonstrate that the majority of blood and saliva cf-mtDNA is likely contained within extracellular circulating mitochondria-derived particles.

### 2.3. Saliva and blood cf-mtDNA stress reactivity

Saliva cf-mtDNA showed a significant increase as fast as 5 minutes after the TSST onset (Figure 2A), reaching its peak at 10 minutes with an average 280% increase (g=0.55, p<0.0001). When normalized to each individual’s baseline values to assess the relative change in cf-mtDNA, this corresponded to a 950% increase at 20 minutes. The initial, rapid rise in saliva cf-mtDNA was followed by a recovery phase where the levels only ceased being significantly higher than the baseline at 120 minutes.

**Figure 2.**
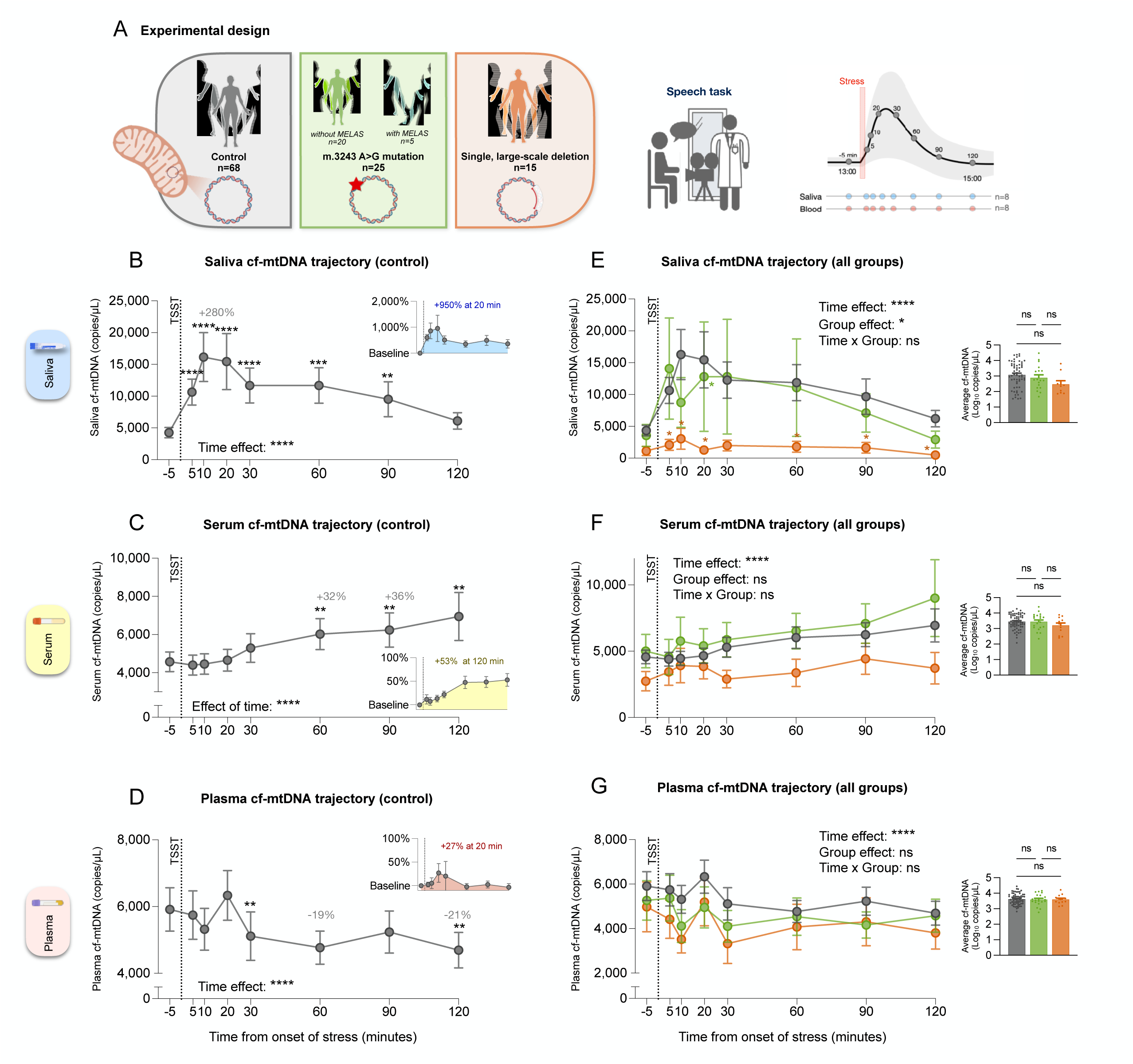
Cell-free mitochondrial DNA reactivity. **(A)** Overview of experimental design. The study included a control group of participants without mitochondrial disease (n=68) and individuals with a mitochondrial genome mutation (m.3243A>G point mutation, both with and without MELAS (Mitochondrial Encephalomyopathy, Lactic acidosis, and Stroke-like episodes); n=25), or a deletion (single, large-scale deletion of mitochondrial DNA; n=15). Participants were exposed to a modified version of the Trier Social Stress Test (TSST) speech task. Saliva, serum, and plasma samples were collected between the hours of 13:00 and 15:00 at 8 timepoints (−5, +5, 10, 20, 30, 60, 90, and 120 minutes relative to stressor onset). Cell-free mitochondrial DNA (cf-mtDNA) trajectories in **(B,E)** saliva (n=68 control, n=23 mutation, n=13 deletion), **(C,F)** serum (n=65 control, n=22 mutation, n=14 deletion), and **(D,G)** plasma (n=65 control, n=22 mutation, n=13 deletion). Dotted line labeled ‘TSST’ indicates the start of the speech task. **(B,C,D)** Average cf-mtDNA trajectories of control participants. Asterisks indicate significant differences between values at indicated timepoints and baseline. Inset plots show average percent changes from individuals’ baseline values (−5 minutes timepoint). **(E,F,G)** Average cf-mtDNA trajectories of participants from all groups. Asterisks indicate significant differences relative to control group value at the corresponding timepoint. Groups are indicated by colors as in (A). **(Inset E,F,G)** Bar graphs comparing individuals’ average cf-mtDNA values over all timepoints from each group. (B-G) Data shown as average ± SEM. Baseline-corrected cf-mtDNA trajectories by study groups are shown in *Figure S5*. Effect sizes and p-values from **(B-G)** mixed-effects analysis of log transformed data with Dunnett’s multiple comparisons test and **(Inset E,F,G)** one-way ANOVA of log transformed data with Tukey’s multiple comparisons test. *p<0.05, **p<0.01, ***p<0.001, ****p<0.0001.

In blood, consistent with previous studies ^10^, serum cf-mtDNA showed a significant increase only after 60 minutes (32%, g=0.20, p<0.01, Figure 2C), but it kept increasing during the rest of the time course (at 90 minutes 36%, g=0.21, p<0.01; at 120 minutes 51%, g=0.19, p<0.01). Unlike our findings for saliva cf-mtDNA, we were unable to capture the onset of a recovery phase for serum cf-mtDNA, preventing us from determining the timing and extent of the maximum reactivity in that biofluid. Finally, plasma cf-mtDNA exhibited a distinct trajectory, initially showing a trend of an increase at 20 minutes (7.1%, ns, Figure 2D), followed by a steady decline that reached significantly lower than baseline levels by 120 minutes (−21%, p<0.01). However, it remains unclear whether this decline stabilized or continued to intensify over time.

Comparing saliva to blood cf-mtDNA reactivity, saliva-based reactivity was on average ∼20-fold larger than in serum (at +60 min; p<0.0001) and ∼18-fold larger than in plasma (within +10-30 min; p<0.0001) (Figure S2A). The magnitudes of individuals’ normalized cf-mtDNA reactivities in different sample types were not correlated (Figure S2B; p>0.05 for all comparisons), again reflecting their independent regulation.

As mentioned above, cf-mtDNA can be released in the circulation via both active release mechanisms (e.g. vesicles) and possibly cell death, while cf-nDNA is believed to only be released after cell death ^9,25–28^. This makes the mitochondrial:nuclear ratio (cf-mtDNA/cf-nDNA) potentially meaningful to isolate the contribution of mitochondrial release. Since cf-nDNA was not detected in most of the plasma samples, we could not compute the cf-mtDNA/cf-nDNA ratio in this biofluid. Cf-mtDNA/cf-nDNA ratios in saliva were, on average, lower than those in serum (Figure S3A; p<0.0001), consistent with baseline measurements of cf-mtDNA and cf-nDNA in these biofluids. These ratios in saliva and serum were not correlated (Figure S3B). Saliva cf-nDNA trajectories showed a significant but smaller effect of time compared to cf-mtDNA trajectories (Figure S1E,F p<0.05). Additionally, we found that saliva cf-mtDNA/cf-nDNA trajectories mirrored cf-mtDNA trajectories, indicating that even after accounting for the co-release of nuclear and mitochondrial genomes, participants still exhibited saliva cf-mtDNA reactivity (Figure S3C,D). On the other hand, serum cf-nDNA and cf-mtDNA/cf-nDNA trajectories did not show a significant time effect (Figure S1E,F, S3C,D).

### 2.4. cf-mtDNA reactivity in genetic mitochondrial diseases

To determine if systemic mitochondrial health regulates blood and saliva cf-mtDNA stress reactivity, as part of the MiSBIE study^3^ we exposed to the same socio-evaluative challenge individuals with rare genetic defects in the mitochondrial genome causing MitoD (n=40, mean age 39 years, 67.5% women). We studied two different groups of individuals presenting either: 1) the 3243A>G mtDNA mutation (n=25); or 2) a single, large-scale mtDNA deletion (n=15), where a segment of the mitochondrial genome is deleted (Figure 2A). Both conditions affect mitochondrial energy transformation and cause multisystem diseases ^29,30^. Baseline cf-mtDNA values were not significantly different between the groups in any sample type (Figure S4A-C).

Although no group differences were observed in baseline saliva cf-mtDNA levels (Figure S4A), over the time course saliva cf-mtDNA trajectories were significantly different between groups (p=0.026; Figure 2E, the baseline adjusted trajectories are shown in Figure S5A). Like controls, MitoD participants also exhibited an acute stress reactivity in saliva cf-mtDNA. On average, relative to controls, absolute cf-mtDNA reactivity in saliva was 10% lower in the mutation group and 530% lower in the deletion group (Figure S4D). In absolute terms, cf-mtDNA increased on average (at +5-10 min) in the control, mutation, and deletion groups by 15,205, 13,941, and 2,414 copies per µl, quantifying the blunted response in the MitoD deletion group, and suggesting that intrinsic mitochondrial health influences saliva cf-mtDNA reactivity (Figure S4D). Saliva cf-nDNA and cf-mtDNA/cf-nDNA average values and trajectories (Figure S1G-I, S3E-G) did not differ by groups.

In serum and plasma, cf-mtDNA trajectories and reactivity (both absolute and normalized) did not differ between groups (Figure 2F,G, Figure S4E,F, Figure S5B,C). Absolute cf-mtDNA reactivity in plasma (at +10-30 min) showed a trend to be lower in mutation (85% less, ns) and deletion groups (80% less, ns) compared to controls (Figure S4F). Serum cf-nDNA (Figure S1G,I,J) and cf-mtDNA/cf-nDNA average values and trajectories (Figure S3E,G,H) did not differ by groups. The correlations between baseline cf-mtDNA values (Figure S4G,H) and normalized cf-mtDNA reactivity (Figure S4I,J) across biofluids in mitoD groups did not differ from the trends found in controls.

## 3. Discussion

By combining experimental repeated-measures stress paradigms and molecular quantification of mtDNA in both saliva and blood, we discovered that cf-mtDNA is rapidly and robustly induced in human saliva, and that it follows kinetics and a pattern distinct from blood collected simultaneously from the same individual. The physiological function or purpose of stress-induced cf-mtDNA reactivity is unknown. The electron microscopy imaging of biofluid fractions enriched for cf-mtDNA demonstrate that as suspected from previous work ^4^, human biofluids including saliva contain mitochondria-related vesicular material, raising the possibility that the majority of cf-mtDNA measured in this and previous studies are in fact circulating mitochondria or mitochondrial fragments, rather than naked DNA.

Our results contrasts with a recent study of saliva cf-mtDNA stress reactivity ^21^. The study by Limberg et al. (2025) reported a trend for a 50% (g=0.14, ns, n=43) increase in saliva cf-mtDNA at 15 minutes post stress. While the timing of the response was similar in both studies finding peak cf-mtDNA reactivity around 10-15 minutes, the magnitude of cf-mtDNA reactivity was considerably larger in our study with a 280% increase (g=0.55, p<0.0001, n=60). This difference could arise from methodological and technical differences. The study by Limberg et al. (2025) employed a modified TSST, consisting of a mock job interview and an arithmetic test, and sessions were conducted during the COVID-19 pandemic during which the interviewer wore a mask (hiding potentially relevant facial features). This setup may have resulted in a less intense stressor than our paradigm, where participants had to prepare and deliver a speech defending themselves against an alleged theft in front of a camera, a mirror, and an evaluator providing neutral to negative feedback. Notably, in the MiSBIE study, only the latter 60% of participants encountered a masked evaluator due to regulatory restrictions, which may have influenced stressor intensity to a lesser extent. Technically, the centrifugation speeds and durations used to extract saliva from the cotton swab were 2.5-fold higher and 200% longer in the Limberg et al. (2025) study than in our study (2,500g x 15 min vs 1,000g x 5 min). Elevated centrifugation speeds create shear force that can damage cells as saliva is extracted from the matrix of the cotton swab, potentially causing artifactually elevated cf-mtDNA levels. Moreover, the cf-mtDNA quantification methods also differed between studies. The process in the Limberg et al. (2025) study does not include any steps between centrifuging samples and qPCR, whereas the method in the current study employed a step to lyse membranes, which we previously showed to recover DNA more uniformly than DNA purification kits ^22^. These methodological differences could introduce biases and inconsistencies in DNA quantification ^31–34^. Thus, while both studies show elevated cf-mtDNA after acute mental stress, differences in stress paradigms, sample processing, and quantification methods could account for the differences in magnitude of this psychobiological response.

The mechanisms of mtDNA release and forms of mtDNA in saliva and blood also remain unclear. Previous studies have shown different mechanisms by which extracellular mitochondria or mitochondrial DNA can originate ^35^ (e.g. release of mtDNA from activated platelets in serum ^26^), and various extracellular bodies that cf-mtDNA may represent ^4–9^. Additionally, our electron micrograph results show that the cf-mtDNA measured in samples from this study is likely a combination of different species of circulating membrane-bound bodies that contain mitochondrial material. It is not clear how the mechanisms of release and species of extracellular vesicles that account for cf-mtDNA dynamics in blood and saliva in response to acute psychological stress relate to those that determine cf-mtDNA levels in response to other factors, such as exercise or chronic mental or physical health conditions. Future studies aiming to use mtDNA as a biomarker should investigate the different circulating forms to increase biological specificity, using different centrifugation speeds or via other physical methods to separate different mtDNA fractions ^9,19^.

One outstanding question to be resolved in future studies relates to the purpose of cf-mtDNA. Why does a stressed organism release mtDNA or whole mitochondria in saliva? While we do not have a definitive answer, we can speculate on an evolutionary explanation that relates to our animal nature. Cell-free mitochondria are readily internalized by various cell types where they influence cell bioenergetics and optimize certain functions ^8,9,36,37^. In the mouse skin for example, this applies to dermal stem cells where exogenously applied mitochondria are internalized and accelerate wound healing after injury ^36^. Mammals instinctively respond to injury by licking their wounds – i.e., locally applying saliva – generally understood to promote coagulation and have an antimicrobial effect ^38^. If as in our studies, injuries and/or the psychological stress associated with the injurious encounter increase saliva cf-mtDNA and mitochondria, then perhaps licking becomes a mechanism to locally apply mitochondria-rich biofluid to promote the healing process, by mitochondria transfer ^8,9,19,36,37^. Future studies should aim to determine the function(s) of saliva cf-mtDNA in humans under different psychobiological states.

This study includes notable limitations. Participants were only subjected to a single stress visit without a control condition, which would be required to separate the effect of the psychosocial stress task from the effect of the overall intervention (intravenous catheter insertion and/or the novelty of the testing environment). The rarity of MitoD makes it difficult to recruit individuals with genetically consistent diseases. Thus, while the sample size for the mutation and deletion MitoD MiSBIE groups is substantial for this population, the absolute numbers of participants is small, inviting caution in interpreting differences between groups. Technically, our electron microscopy studies emphasize the diversity and heterogeneity of mitochondria-like particles in saliva and blood. As outlined previously ^7,19^, there are several different types of microparticles, mitochondria, platelet-derived fragments, and exosomes in the mtDNA-enriched biofluid fractions. Ideally, each would be biochemically and/or physically isolated and purified, assayed for mtDNA in parallel with protein markers, thus revealing which “type” of particles contain the baseline- and stress-inducible fractions of cf-mtDNA. Technical limitations and the scale of this study precluded us to perform these detailed analyses. Future studies of human cf-mtDNA will benefit from including further levels of analytical specificity and adopting standard nomenclature ^7,19^.

In summary, we have showed that human biofluid cf-mtDNA levels are acutely influenced by a brief socio-evaluative stress. In particular, we show for the first time that saliva cf-mtDNA is robustly stress inducible, calling for further studies examining the usability of saliva cf-mtDNA as a biomarker of acute and/or chronic stress exposure, and potentially a marker of systemic mitochondrial recalibrations. Our studies in rare MitoD patients with objective impairments in mitochondrial health suggested that saliva cf-mtDNA levels could indirectly reflect systemic mitochondrial disorders, although this point requires further research. Together, the inducibility and accessibility of *saliva* cf-mtDNA provides a foundation to examine the psychobiological regulation of cell-free mitochondria in vulnerable populations where collecting blood is challenging. Given the dynamic properties uncovered here, future work should consider intensive repeated-measures studies where saliva is sampled multiple times per day^14^, or even per hour, depending on the question. The large inter-individual differences in baseline saliva cf-mtDNA levels also call for large-scale studies where the origin of this variance can be ascertained, including deeply-phenotyped cohorts with existing saliva biobank repositories.

## 4. Methods

### 4.1. Participants

Participants were enrolled in the MiSBIE study ^3^ in adherence to the directives outlined by the New York State Psychiatric Institute IRB protocol #7424 and the Columbia University Medical Center IRB protocol #AAAU9470. The study was registered in <u>ClinicalTrials.gov</u> under #NCT04831424. Recruitment was conducted both within our local clinic at the Columbia University Irving Medical Center, and nationally throughout the United States and Canada. All enrolled participants provided written informed consent, authorizing their participation in the investigative procedures and the dissemination of findings. Recruitment occurred from June 2018 to May 2023, including a total of 68 healthy controls (66% women) with blood and data for the acute stress protocol, 25 mitochondrial disease (MitoD) patients presenting the 3243A>G mutation (64% women) and 15 with a single large-scale mtDNA deletion (73% women).

Inclusion criteria for all groups were 1) ages between 18-60, 2) willing to provide saliva samples and have a venous catheter installed for blood collection, 3) willing to provide informed consent and having the capacity to consent, 4) use of effective birth control method for women of childbearing capacity, and 5) English speaking. Exclusion criteria for all groups were 1) cognitive deficits that prohibited providing informed consent, 2) symptoms of flu or seasonal infection within four weeks preceding visit, 3) Raynaud’s syndrome, 4) involvement in therapeutic trials listed on clinicaltrials.gov, including exercise, 5) having metal in body or claustrophobia that would prevent MRI. Additional exclusion criteria for the control group was a diagnosis of mitochondrial 3243A>G mutation or mitochondrial DNA deletion.

Inclusion criteria for the MitoD deletion and mutation without MELAS (Mitochondrial Encephalomyopathy, Lactic Acidosis, and Stroke-like episodes) groups also included 1) having a single large scale DNA deletion or m.3243A>G point mutation and 2) taking a confirmatory genetic test or consenting to undergo genetic testing. Exclusion criteria for those groups included 1) neoplastic disease, 2) history of strokes and seizures, 3) involvement in therapeutic trials listed on clinicaltrials.gov, including exercise, and 4) clinical use of steroid therapy. Having metal in body preventing MRI was not an exclusion criterion for this group.

Inclusion and exclusion criteria for the MitoD mutation with MELAS group were the same as those for the mitochondrial deletion and mutation without MELAS groups, except the inclusion criteria also required that participants have had at least one stroke-like episode, seizure, or both, in addition to having the m.3243A>G mutation, and the exclusion criteria did not include history of strokes and seizures.

### 4.2. Stress Reactivity Protocol

A psychosocial stress response was elicited in participants using the TSST as described in full detail in ^3^. Participants were seated in a room. Five minutes after baseline samples and readings were taken, the study coordinator entered the room with a clipboard and video camera, which was set up to appear as if it was recording the participant. A full-length mirror was also set up so participants can see their reflection. The test coordinator instructed participants to prepare a speech to defend themselves from an accusation of stealing store merchandise. After the instructions, participants were given 2 minutes to prepare their speech. A person wearing a lab coat posing as an ‘evaluator’ stood in the room and took notes using a clipboard as participants delivered their speech for 3 minutes.

### 4.3. Blood and saliva collection and processing

Blood and saliva were collected at 5 minutes prior to the start of the stress task and at 5, 10, 20, 30, 60, 90, and 120 minutes after the start of the stress task. For blood collection, participants were fitted with an intravenous catheter in the right arm >45 minutes prior to the start of the stress task.

Serum and plasma were collected in red-top (BD, REF367815) and K2 EDTA (BD REF367899) vacutainers respectively. Serum tubes were inverted 10-12 times upon collection and incubated for 30 minutes at room temperature. They were then centrifuged at 2,000 x g for 3.5 minutes at room temperature then transferred to ice in a Styrofoam container. Plasma tubes were processed identically to serum tubes except they were not incubated for 30 minutes before centrifugation. Plasma and serum tubes were transported to the laboratory (∼5 minute walk) and centrifuged at 2,000 x g for 10 minutes at 4 °C. ∼80% of the supernatant from each tube was transferred to a new 15 mL tube and centrifuged at 2,000 x g for 10 minutes at 4 °C. ∼90% of this supernatant was transferred to a new 15 mL tube, aliquoted into three or more 2 mL cryovials and stored at −80 °C. Samples were later thawed, aliquoted into 96-well plates, and re-frozen in preparation for plate-based analyses. Blood was not available from ten participants due to phlebotomy failure.

Saliva was collected using salivettes (Starstedt Cat# 51.1534.500). Participants were instructed to hold a cotton salivette swab on the center of the tongue for 2-5 minutes without moving or biting it. After two minutes, the study coordinator would check the swab to determine if it was saturated or if it required more time. After collection, swabs were immediately returned to plastic tubes and kept on ice in a Styrofoam container, then transported to the lab where they were centrifuged at 1,000 x g for 5 minutes at 4 °C. Supernatants were aliquoted into 2 mL cryovials and stored at −80 °C. The samples were later thawed and centrifuged at 5,000 x g for 10 minutes at room temperature. The resulting 5,000 x g supernatants were aliquoted into 96-well plates and stored at −80 °C until analysis.

### 4.4. cf-mtDNA detection

mtDNA (represented by mitochondrial gene *ND1*) and nDNA (represented by nuclear gene *B2M*) in cell-free blood and saliva samples were quantified using previously published methods^22^. Briefly, samples were diluted 20-fold in freshly assembled lysis buffer (Tris-HCl, 114 mM, pH 8.5; Tween-20, 6%; Proteinase K, 0.2 mg/ml) on two replicate 96-well plates and heated to 55 °C for 10 hours for thermolysis, then heated to 95 °C for 10 minutes to deactivate Proteinase K and other enzymes. Lysates were held at 4°C and processed further the following day or stored at −80 °C if processing did not occur immediately. 8 µL of each replicate lysate was transferred to three wells of a 384-well plate that had been pre-loaded with 12 µL of freshly assembled qPCR buffer (1x TaqMan Universal MasterMix Fast; 0.3 µM *ND1* primers; 0.1 µM *ND1* probe; 0.3 µM *B2M* primers; 0.1 µM *B2M* probe). An eight-step 1:4 serial dilution of DNA extracted from a human fibroblast culture (hFB1) that had been quantified using digital PCR was also added to each 384-well plate in triplicate as a standard curve. TaqMan-based qPCR reactions were run and measured using a QuantStudio 7 Flex system. For each replicate sample, the copy number of each target gene per µL was determined by comparing the median of its three Ct values for that gene against the median Cts measured for the standard curve on the same 384-well plate and accounting for dilution into lysis and qPCR buffers. Reactions with Cts representing three or fewer copies of target gene per µL were considered below the limit of detection (<LOD) and omitted from the analysis (Saliva: 3.5% of *ND1* values and 49.5% of *B2M* values <LOD. Serum: 0.1% of *ND1* values and 51.6% of *B2M* values <LOD. Plasma: 0.0% of *ND1* values and 98.7% of *B2M* values <LOD). The scarcity of nDNA in our samples may be attributed to 1) the specific release of membrane-bound particles containing mtDNA, but not nDNA, into circulation, and 2) the relative abundance of mtDNA over nDNA on a cellular basis. For each sample, the copy numbers determined for each replicate were averaged to yield a single value. Replicate plates were re-run starting from lysis if the median coefficient of variance between replicate values was greater than 4%.

### 4.5. Data processing and statistical analysis

Individual participants’ absolute cf-mtDNA reactivities were defined as the difference between their baseline value and the maximum value within a range of time points for saliva (+5-10 minutes after the onset of the stress task) and plasma (+10-30 minutes after the start of the task), and at +60 minutes in serum. Participants’ normalized reactivities for each sample type were defined as absolute reactivity divided by baseline and are presented as a percentage. Statistical tests were performed using GraphPad Prism version 10.4.0 (GraphPad Software, Boston, Massachusetts USA). Because cf-mtDNA and cf-nDNA values were found to follow a lognormal distribution, values were log transformed before statistical analyses. Welch’s t tests and one-way ANOVA with Tukey’s multiple comparisons test were used to compare averages and baseline values between two and three groups, respectively. Effect sizes are presented as bias corrected Hedges’ *g*. cf-mtDNA reactivities (both absolute and normalized) included negative values that could not be log transformed, so Kruskal-Wallis tests with Dunn’s test were used to compare these values between groups and sample types. Correlations between individuals’ cf-mtDNA baseline values and normalized reactivities in different sample types were computed using Spearman correlation. Mixed-effects model ANOVA was used to evaluate the effects of time and participant group on cf-mtDNA and cf-nDNA measurements, with Dunnett’s multiple comparisons test to compare average values at baseline with those measured at subsequent time points and values in mitochondrial mutation and deletion groups with those from the control group.

## Acknowledgements

The MISBIE study was supported by several sources, including National Institute of Health (NIH) grants R21MH113011, R01MH122706, RF1AG076821, and R01MH137190 to M.P. Additional support came from the Seed Grant Program for MR Studies at Columbia University’s Zuckerman Mind Brain Behavior Institute, the Robert N. Butler Columbia Aging Center Fellowship Program at the Mailman School of Public Health, the National Center for Advancing Translational Sciences, and NIH grants UL1TR001873 and P30CA013696. The study also benefited from support from the Columbia Irving Institute Scholars Program, the Wharton Fund, and the Baszucki Group to M.P.

## Data availability

Data will be made available upon request.

## Author contributions

M.P., R-P.J. and C.T. designed the study. J.S. and T.Y. performed the cf-mtDNA assays. J.S. C.T. and D.S. ran the statistical analysis. C.T. and D.S. wrote the manuscript. All authors reviewed the final version of the manuscript.

