## Supplemental Figures for "Saliva and Blood Cell-Free mtDNA Reactivity to Acute Psychosocial Stress"

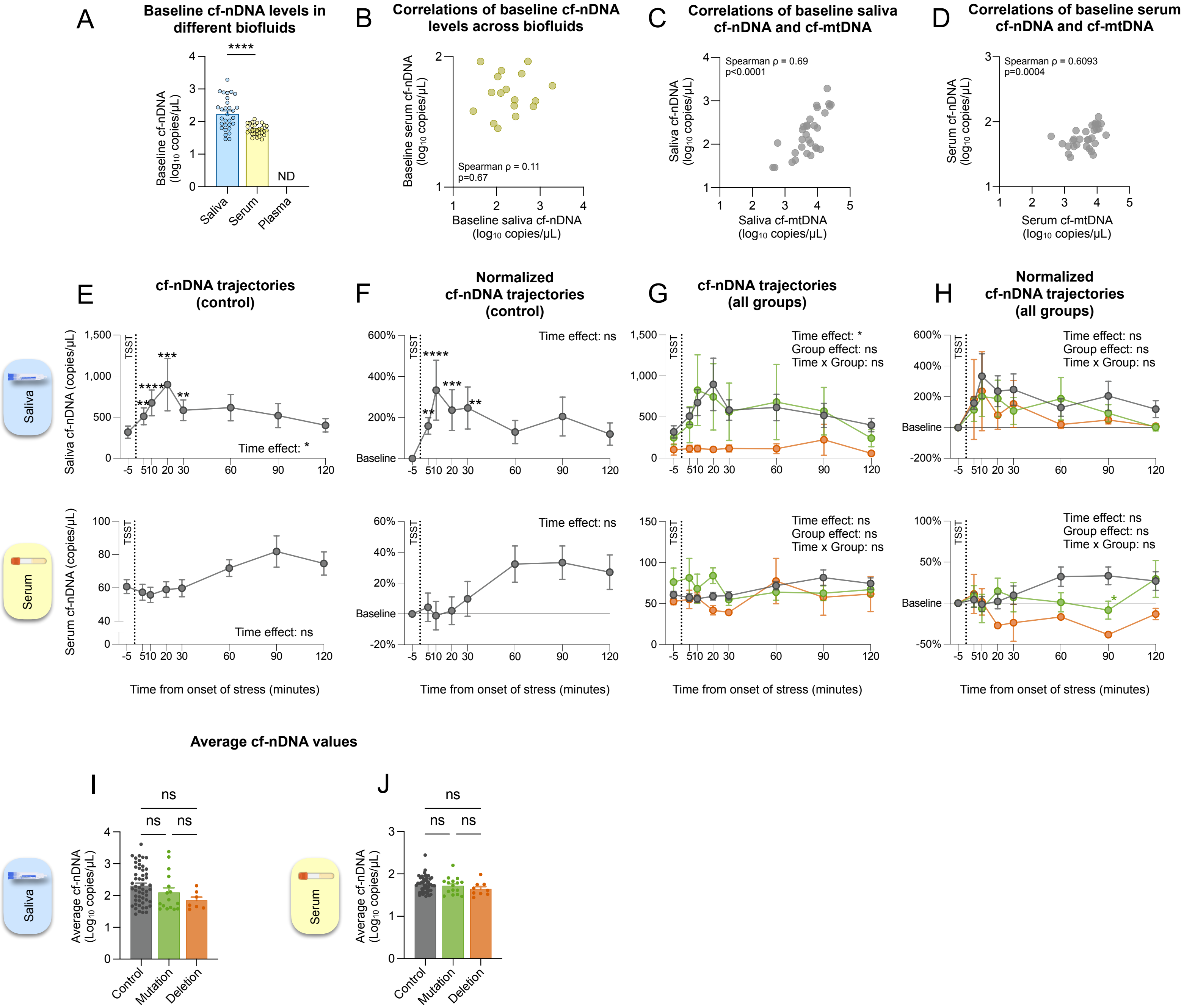


**Supplemental Figure 1. Cell-free nuclear DNA reactivity. (A)** Comparison of control group cf-nDNA levels at baseline in saliva and serum. Plasma cf-nDNA was detected in only a single baseline sample (not shown). **(B)** Scatter plot illustrating relation between control group baseline cf-nDNA levels in saliva and serum. Only 17 control group participants had baseline samples for both saliva and serum with B2M values above LOD. **(C,D)** Scatter plots illustrating relation between cf-mtDNA and cf-nDNA measurements in control group baseline saliva (n=30) and serum samples (n=30) for which both values were available. **(E-H)** cf-nDNA trajectories in saliva (n=76) and serum (n=76). Dotted line labeled ‘TSST’ indicates the start of the speech task. Data shown as average ± SEM. cf-nDNA was detectable in only 1.3% of plasma samples; plasma trajectories not plotted. **(E,G)** Average cf-nDNA trajectories in saliva (n=53 control, n=16 mutation, n=7 deletion) and serum (n=51 control, n=16 mutation, n=9 deletion). Asterisks indicate significant differences between values at indicated timepoints and baseline. **(F,H)** Same as (E,G) with values normalized to individuals’ baselines (-5 minute timepoint) before averaging. Asterisks in (F) indicate significant difference between values at indicated timepoints and baseline. Asterisks in (H) represent significant difference between indicated group and control group at indicated timepoints. **(I,J)** Average cf-nDNA values measured over all timepoints in saliva and serum for individuals in the control (saliva n=53; serum n=51), mutation (saliva n=16; serum n=16) and deletion (saliva n=7; serum n=9) groups. Effect sizes and p-values from **(A)** Welch’s t test, **(B-D)** Spearman rank correlation, **(E-H)** mixed-effects model of log transformed values with Dunnett’s multiple comparisons test, and **(I,J)** one-way ANOVA with Tukey’s multiple comparisons test. *p<0.05, **p<0.01, ***p<0.001, ****p<0.0001.

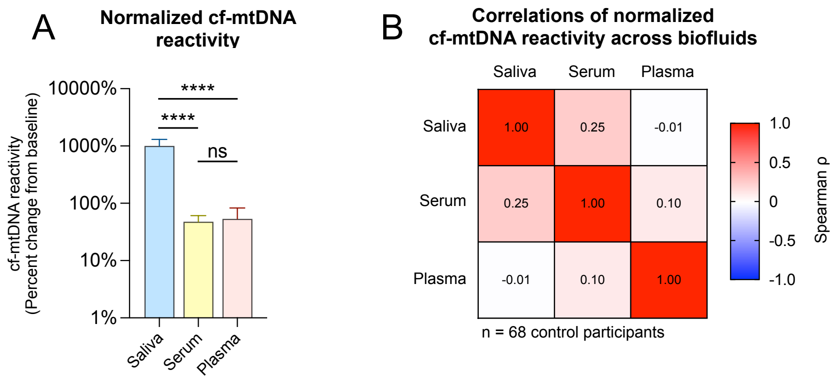


**Supplemental Figure 2. cf-mtDNA reactivity across biofluids. (A)** Comparison of normalized cf-mtDNA reactivity in saliva, serum, and plasma in control participants, where reactivity is defined as the percent difference between baseline and the maximum value measured between 5-10 minutes for saliva, the value at 60 minutes for serum, and the maximum value between 10-30 minutes for plasma (saliva n=57, serum n=61, plasma n=63). **(B)** Heatmap illustrating correlations (Spearman ρ) of normalized cf-mtDNA reactivity (as defined in (A)) between sample types for healthy controls. (A) Bars represent average ± SEM. Effect sizes and p-values from **(A)** Kruskal-Wallis test with Dunn's multiple comparisons test and **(B)** Spearman rank correlation. *p<0.05, **p<0.01, ***p<0.001, ****p<0.0001.

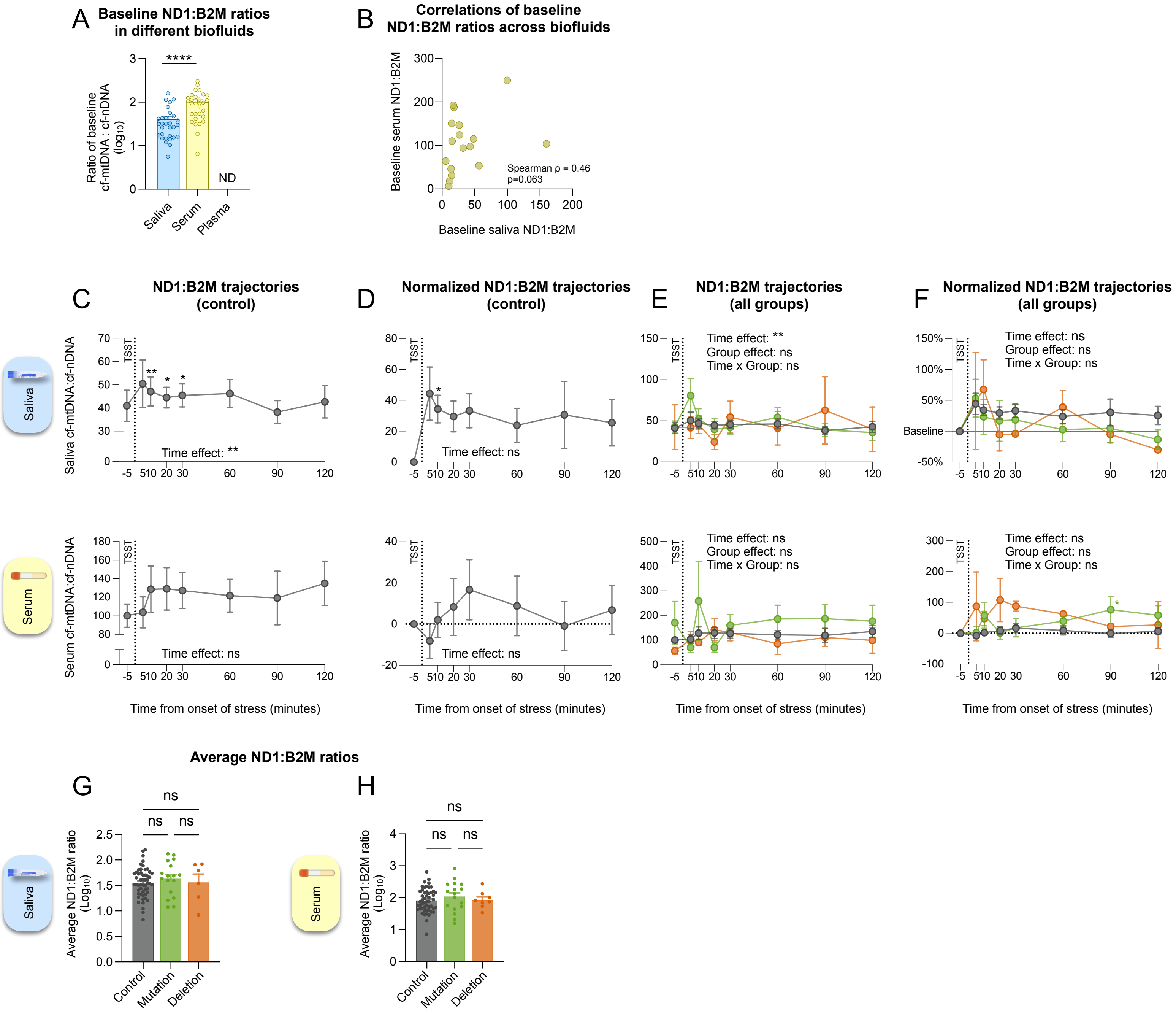


**Supplemental Figure 3. Cell-free mtDNA : nDNA ratio reactivity. (A)** Comparison of baseline cf-mtDNA : cf-nDNA ratios in saliva and serum (saliva n=30, serum n=30). Plasma cf-nDNA : cf-mtDNA could be calculated in only a single baseline sample (not shown). **(B)** Scatter plot illustrating relation between cf-mtDNA : cf-nDNA ratio in saliva and serum (n=17, only this subset of control group participants had baseline samples for both saliva and serum with B2M values above LOD). **(C-F)** Trajectories of cf-mtDNA : cf-nDNA ratios in saliva (n=53 control, n=16 mutation, n=7 deletion) and serum (n=51 control, n=16 mutation, n=9 deletion) shown as average ± SEM. cf-nDNA was detectable in only 1.3% of plasma samples; plasma cf-nDNA : cf-mtDNA ratio trajectories not plotted. **(C,E)** cf-mtDNA : cf-nDNA ratios for control and all groups. **(D,F)** Same as (C,E) with average percent change in ratio relative to baseline (-5 minute timepoint). **(G,H)** Average cf-mtDNA : cf-nDNA ratios in saliva and serum over all timepoints for each group (saliva n=76, serum n=76). Effect sizes and p-values from **(A)** Welch's t test, **(B)** Spearman rank correlation, **(C-F)** mixed-effects model of log transformed values with Dunnett's multiple comparisons test, and **(G,H)** one-way ANOVA with Tukey's multiple comparisons test. *p<0.05, **p<0.01, ***p<0.001, ****p<0.0001.

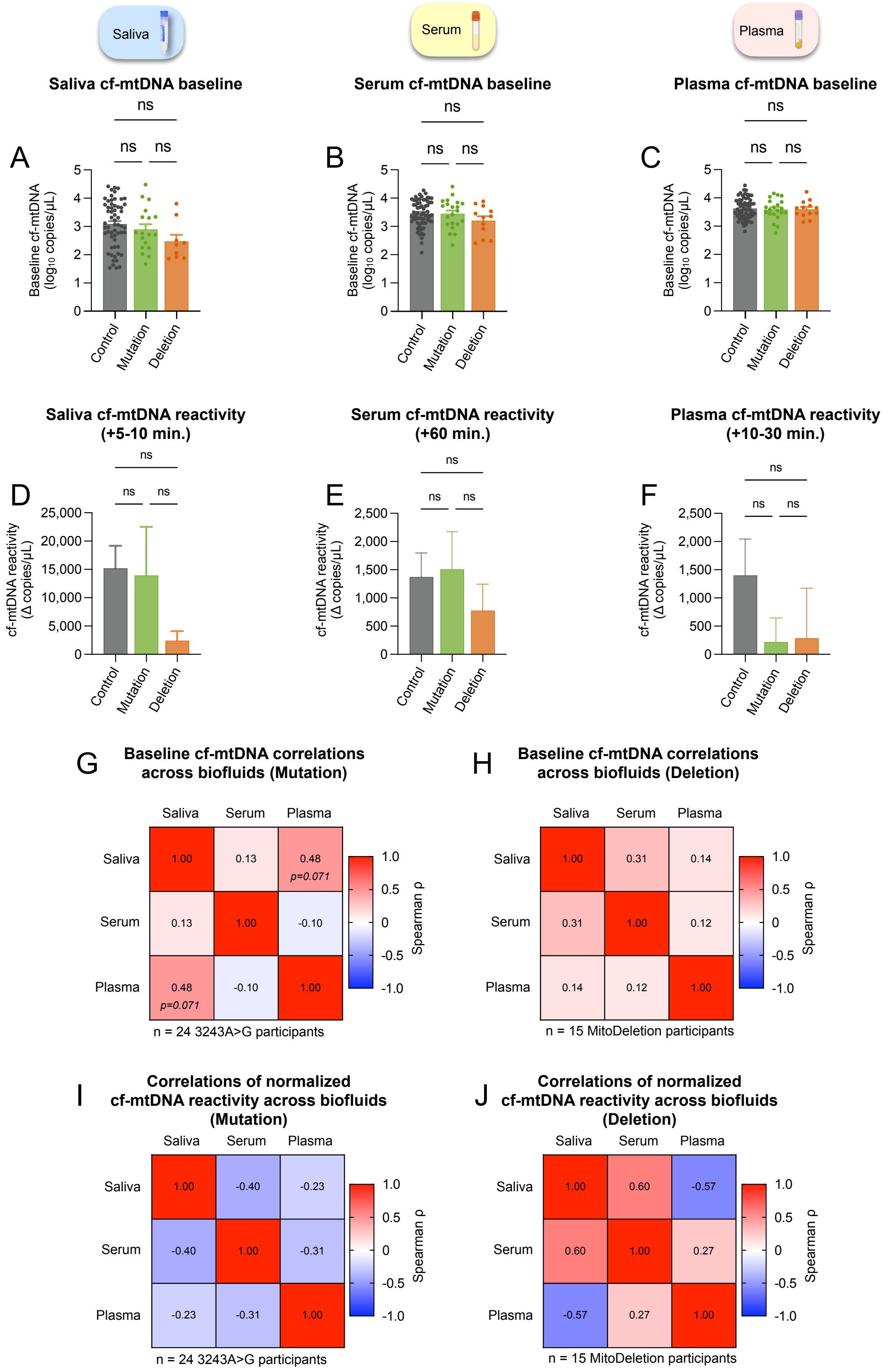


**Supplemental Figure 4. cf-mtDNA baseline and reactivity in controls vs mitoD.** Comparisons of average cf-mtDNA baseline values between study groups for **(A)** saliva (n=59 control, n=18 mutation, n=9 deletion) **(B)** serum (n=65 control, n=21 mutation, n=12 deletion) and **(C)** plasma (n=64 control, n=21 mutation, n=13 deletion). **(D-F)** Same as in (A-C) for absolute cf-mtDNA reactivity, defined as the difference between baseline and the maximum value measured from 5-10 minutes for saliva (n=57 control, n=18 mutation, n=9 deletion), the value at 60 minutes for serum (n=61 control, n=21 mutation, n=9 deletion), and the maximum value from 10-30 minutes for plasma (n=63 control, n=21 mutation, n=13 deletion). Heatmaps illustrating correlations (Spearman ρ) between baseline cf-mtDNA levels in saliva, serum, and plasma for participants **(G)** with 3243A>G mutation and **(H)** with a single large-scale mitochondrial deletion. **(I, J)** Same as in (G,H) for normalized cf-mtDNA reactivity, defined as percent change between baseline and sample-type specific time ranges defined in (D-F). Effect sizes and p-values from (A-C) one-way ANOVA with Tukey's multiple comparisons test, (D-F) Kruskal-Wallis test with Dunn's multiple comparisons test, (G-J) spearman rank correlation, *p<0.05, **p<0.01, ***p<0.001, ****p<0.0001.

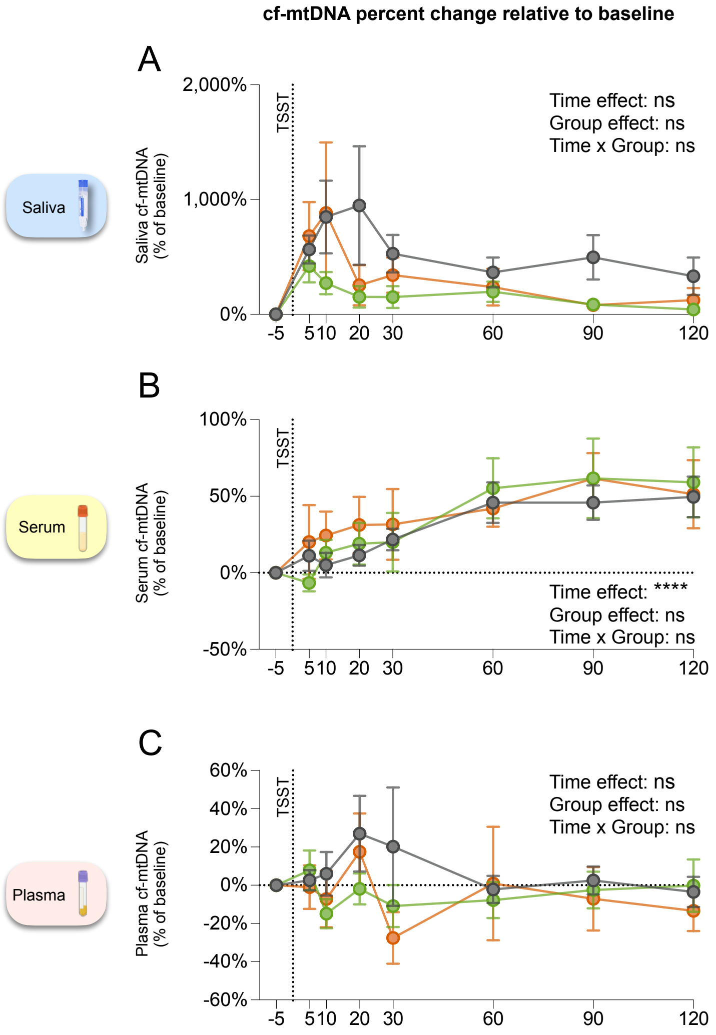


**Supplemental Figure 5. Baseline-corrected cf-mtDNA trajectories. (A-C)** cf-mtDNA trajectories in saliva (n=68 control, n=23 mutation, n=13 deletion) **(A)**, serum (n=65 control, n=22 mutation, n=14 deletion) **(B)**, and plasma (n=65 control, n=22 mutation, n=13 deletion) **(C)** as average percent change relative to baseline (-5 minutes timepoint). Dotted line labeled 'TSST' indicates the start of the speech task. Data shown as average ± SEM. Effect sizes and p-values from **(A-C)** mixed-effects analysis of log transformed data with Dunnett's multiple comparisons test,
